# A randomized controlled trial of antibody response to 2019-20 cell-based inactivated and egg-based live attenuated influenza vaccines in children and young adults

**DOI:** 10.1101/2021.09.02.21263043

**Authors:** Katherine V. Williams, Bo Zhai, John F. Alcorn, Mary Patricia Nowalk, Min Z. Levine, Sara S. Kim, Brendan Flannery, Krissy Moehling Geffel, Amanda Jaber Merranko, Jennifer P. Nagg, Mark Collins, Michael Susick, Karen S. Clarke, Richard K. Zimmerman, Judith M. Martin

**Author notes:** **Corresponding author:** Department of Pediatrics, General Academic Pediatrics, 3520 Fifth Ave., Pittsburgh, PA, USA. (JM Martin). **Additional author emails:** KVW; BZ; JFA; MPN; MZL; SSK; BF; KMG; AJM; JPN; MC; MS; KSC; RKZ.

## Abstract

**Background:** Hemagglutination inhibition (HAI) titers to the live-attenuated influenza vaccine (LAIV4) are typically lower than its counterpart egg-based inactivated influenza vaccines (IIV). Similar comparisons have not been made between LAIV4 and the 4-strain, cell-culture inactivated influenza vaccine (ccIIV4). We compared healthy children and young adult HAI titers against the 2019-2020 LAIV4 and ccIIV4.

**Methods:** Participants aged 4-21 years were randomized 1:1 to receive ccIIV4 (n =100) or LAIV4 (n=98). Blood was drawn prevaccination and on day 28 (21-35) post vaccination. HAI assays against egg-grown A/H1N1, A/H3N2, both vaccine B strains and cell-grown A/H3N2 antigens were conducted. Outcomes were geometric mean titers (GMT) and geometric mean fold rise (GMFR) in titers.

**Results:** GMTs to A/H1N1, A/H3N2 and B/Victoria increased following both ccIIV and LAIV and to B/Yamagata following ccIIV (p<0.05). The GMFR range was 2.4-3.0 times higher for ccIIV4 than for LAIV4 (p<0.001). Within vaccine types, egg-grown A/H3N2 GMTs were higher (p<0.05) than cell-grown GMTs [ccIIV4 day 28: egg=205 (95% CI: 178-237); cell=136 (95% CI:113-165); LAIV4 day 28: egg=96 (95% CI: 83-112); cell=63 (95% CI: 58-74)]. The GMFR to A/H3N2 cell-grown and egg-grown antigens were similar. Pre-vaccination titers inversely predicted GMFR.

**Conclusion:** The HAI response to ccIIV4 was greater than LAIV4 in this study of mostly older children, and day 0 HAI titers inversely predicted GMFR for both vaccines. For both vaccines, the A/H3N2 cell-grown antigen levels were lower than egg-grown, but the GMFR for cell-grown and egg-grown did not differ significantly within vaccine type.

**Clinical Trials No:** NCT03982069

## 1. Introduction

Influenza is a major public health burden causing millions of illnesses and outpatient visits and resulting in tens of thousands of hospitalizations and deaths in the U.S. annually [1]. Vaccination is the primary influenza prevention method. Advances in vaccine technology have resulted in multiple vaccine types licensed for use in the U.S., comprising intramuscularly administered inactivated influenza vaccines (IIV), including a quadrivalent cell culture-based inactivated vaccine (ccIIV4), and nasally administered, egg-based live attenuated influenza vaccine (LAIV4). The Advisory Committee on Immunization Practice (ACIP), which sets civilian immunization policy for the United States, recommends vaccination with any licensed vaccine, without preference [2].

Several factors can affect the immune response to either vaccine type, including pre-existing antibodies, prior vaccination status, and age [3-5]. Nasally administered LAIV4, which is absorbed through the nasal mucosa, generates an immune response that differs from direct systemic exposure from intramuscularly administered IIV [6]. Moreover, the manufacturing process may have an impact on immune response. For example, the viruses used for vaccine antigens can be grown in mammalian cell culture or embryonated chicken eggs. Growth in cell culture has the advantage of limiting the virus mutations that often occur during the egg-based manufacturing process. While, egg-based LAIV4 has been shown to be more effective against antigenically different or drifted strains [7], egg-induced mutations in influenza A(H3N2) vaccine viruses have resulted in reduced efficacy of egg-based vaccines [8].

Previous research comparing the hemagglutination inhibition (HAI) antibody responses of IIV and LAIV4 has historically demonstrated lower HAI titers in response to LAIV compared to egg-based IIV [3, 6, 9, 10]. These studies were conducted before the reformulation of LAIV4 necessitated by poor vaccine effectiveness [11]. In a previous study among children, we found no differences in seroconversion in children receiving egg- or cell-grown IIV in 2018-19 [12]. The cell-grown IIV4 used in that study contained A/H1N1 that was derived from an egg-based seed leading to a 3:1 cell to egg formulation. The 2019-20 season is the first in which the ccIIV4 formulation is exclusively cell-based. Furthermore, the current LAIV4 has been reformulated to avoid the egg-induced mutations to the A/H3N2 antigen. Thus, a comparison of the HAI response to egg- and cell-based antigens for each vaccine in a clinical setting of children and young adults is warranted.

The purpose of this randomized controlled trial was to quantify and compare pre- and post vaccination HAI responses to 2019-20 LAIV4 and ccIIV4, among a racially diverse group of children and young adults during the 2019-20 influenza season.

## 2. Methods

The Institutional Review Boards at the University of Pittsburgh and the Centers for Disease Control and Prevention (CDC) approved this study. Written informed consent and assent, where appropriate, were obtained from all participants and/or their parents/legal guardians prior to beginning study procedures.

### 2.1 Study Design and Participants

This study was a randomized controlled clinical trial comparing the serologic antibody response to two quadrivalent 2019-20 influenza vaccines: ccIIV4 (Seqirus Flucelvax®, lot 261199, 261203) administered intramuscularly in the upper arm deltoid muscle with a 25-gauge 1 inch needle and egg-based LAIV4 (AstraZeneca FluMist®, lot LJ2514) administered intranasally with one metered spray in each nostril.

Healthy participants included patients ages 4-17 years who receive primary care at pediatric and family medicine clinics. These children typically receive annual IV [13] and thus tend to have higher pre-vaccination HAI titers. Other participants up to age 21 who were unvaccinated in the prior season (2018-19) and thus would be expected to have lower pre-vaccination HAI titers, also participated. Participants were recruited and enrolled from September 20, 2019 through November 13, 2019 through: 1) mailings to past research participants; 2) personal approach by research assistants/nurses at well-child visits at five primary care health centers (one pediatric and four family medicine practices); and 3) targeted emails to 18-21-year-olds enrolled in an institutional participant-centered research registry. Recruitment ended when the targeted number for enrollment goal was achieved. All study vaccinations were completed by November 25, 2019, prior to regional circulation of influenza virus; the final participant visit was completed on December 18, 2019. Verification of prior year vaccination status was confirmed through the Pennsylvania Statewide Immunization Information System (PA-SIIS) vaccine registry.

Eligibility criteria included parental consent for randomization to receive one of the two 2019-20 FDA approved influenza vaccines used in the study. Exclusion criteria included: weight <16.8 kg (37 lb.); known to be pregnant; an immunosuppressing health condition or taking immunosuppressant medications; having already received the 2019-20 influenza vaccine; not able to complete all study visits in the appropriate time window; contraindications to LAIV4 (asthma with treatment for wheezing in the past year, individuals who may have contact with severely immunosuppressed persons) or conditions with precautions to LAIV4 vaccine (disorders of the cardiovascular system, seizures, diabetes mellitus, renal or hepatic disease) or severe allergies to eggs or influenza vaccine components.

Following screening and consent, a blood sample was drawn. Participants were then randomized to receive one of the two study vaccines in an unblinded manner. In order to ensure that roughly the same number of each vaccine types were administered at each of the enrollment sites, participants were randomized in blocks of four. Sequentially numbered vaccine assignment cards were computer generated based on 1:1 randomization for each vaccine type. The next available numbered card was used to instruct the clinical staff which vaccine to administer under standard vaccination protocols. While the participant and research team were not blinded to randomization allocation, the laboratory team was unaware of participants’ group assignment.

### 2.2 Demographic data collection

Baseline data were collected with entry into REDCap™, a secure, online database management system. Baseline demographics included age, and self-identified: sex, race, ethnicity, parental educational status, and health insurance coverage.

### 2.3 Hemagglutination inhibition assay

Whole blood samples were obtained on participants pre-vaccination and 28 days (range 21-35 days) post influenza vaccination in BD Vacutainer™ serum separator tubes with polymer gel/silica activator additive (BD 367989). Tubes were held at room temperature and taken to the processing laboratory within 4 hours of being drawn. Aliquoted serum samples were frozen at −80°Celcius until assayed.

Assays were conducted as previously described [11]. Sera were heat inactivated, tested for nonspecific agglutinins, and adsorbed as needed. Sera were serially diluted 2-fold and incubated with 4 hemagglutination units per 25 µL of virus with erythrocytes for quantification of HAI titers. Turkey erythrocytes were used for the testing of A/H1N1 and B influenza viruses, Guinea pig erythrocytes with 20 mM oseltamivir were used for the testing of A/H3N2 viruses. HAI titer was defined as the reciprocal of the last dilution of serum that completely inhibited hemagglutination. Antibody titers <10 (initial sera dilution) were reported as 5 for analysis. Sera for both vaccine types were tested in HAI assays against egg-grown vaccine strains included in the 2019-20 influenza vaccines (A/Brisbane/02/2018 (H1N1)pdm09, A/Kansas/14/2017 (A/H3N2), B/Colorado/06/2017 (B/Victoria lineage) and B/Phuket/3073/2013 (B/Yamagata lineage), as well as an available cell-grown HAI assay for A/Kansas/14/2017 A/(H3N2).

### 2.4 Statistical analysis

Primary outcome measures were post vaccination geometric mean titers (GMT) and geometric mean fold-rise (GMFR) in titers from day 0 to day 28. Secondary outcomes were defined as an HAI titer either 1: ≥40 or 1: ≥110 and seroconversion, defined as a four-fold rise in titer with day 28 post vaccination titer ≥40.

Based on previous research comparing seroconversion rates for LAIV4 and ccIIV4, it was determined that a sample size of 200 would be needed to achieve an 80% power to detect a significant difference between vaccine types for seroconversion with a moderate effect size of 0.2 [11]. Due to high prevaccination GMT, the primary outcome of this study is reported as GMT and GMFR in titers. A post hoc power calculation of A/H1N1-A/Brisbane day 28 titer outcomes revealed a sample size of 200 had power of 99.8% to detect a difference between vaccine types for day 28 GMT, GMFR in titers and seroconversion.

Summary statistics of dichotomous variables were conducted by vaccine group using chi-square/Fisher exact tests for categorial variables and t-tests for continuous variables. Fold-rise was calculated as the ratio of the postvaccination titer to the prevaccination titer. HAI and fold-rise were log_2_-transformed in order to perform t-tests and linear regressions. GMTs, GMFR, and 95% confidence intervals were calculated using the survey means procedure in SAS. Linear regression was used to examine associations between fold-rise controlling for age, pre-vaccination titers, and prior season (2018-19) vaccination status as previously described [14]. All analytical procedures were performed using SAS® 9.4 (Cary, NC). Statistical significance of two-sided tests was set at type I error (alpha)=0.05.

## 3. Results

Of 234 persons aged 4 to 21 years assessed for eligibility, 16 were excluded and 218 were randomized: 110 received ccIIV4 and 108 LAIV4. After randomization, 20 participants were excluded from the per-protocol analysis: 19 failed to return for post-vaccination specimen collection or were missing paired sera, and one participant in the ccIIV4 group received LAIV4. Per-protocol analysis included 198 individuals: 100 received ccIIV4 and 98 received LAIV4 (Figure 1).

**Figure 1:**
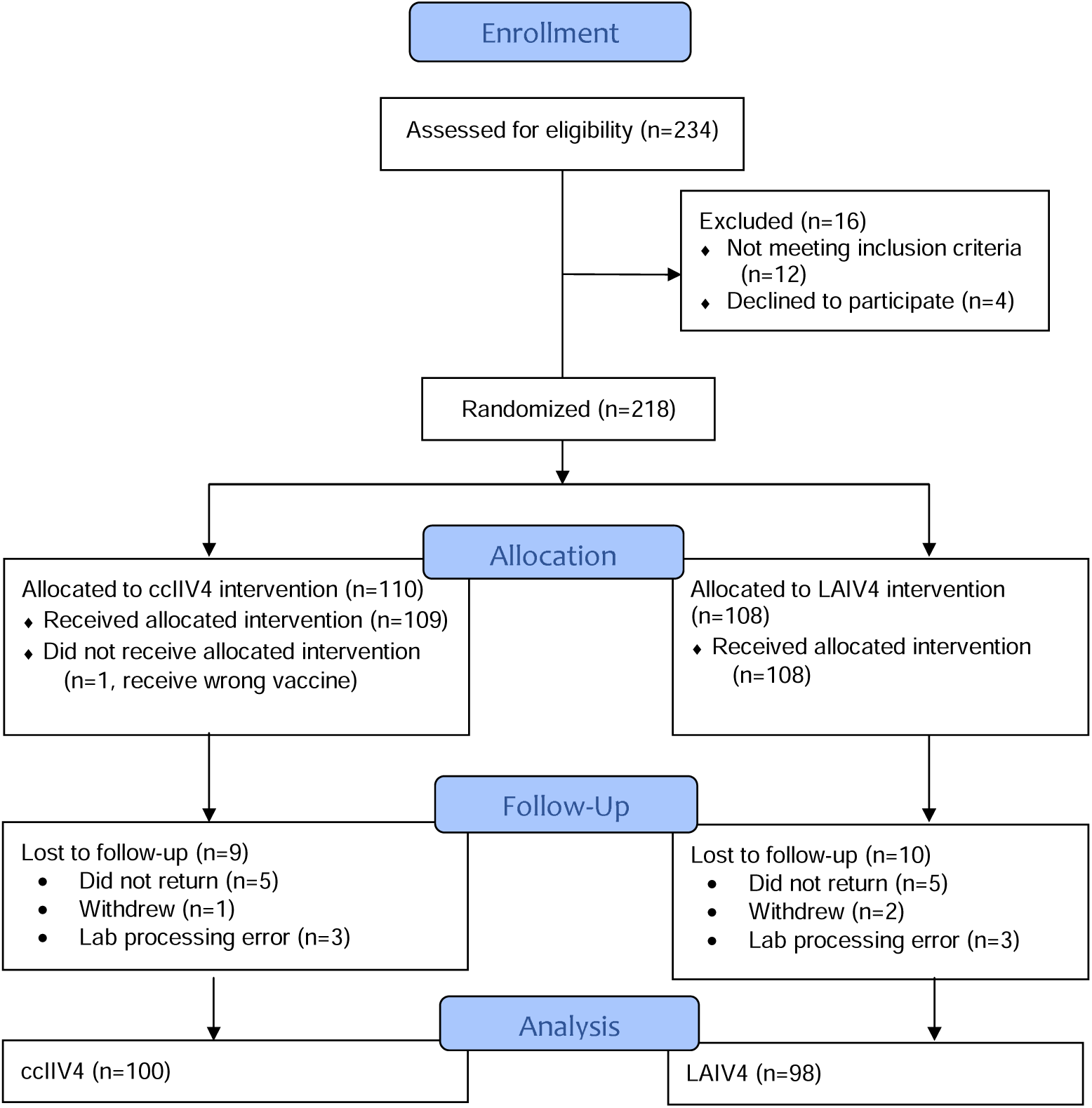
CONSORT 2010 Flow Diagram.

Patient characteristics were similar for the two vaccine arms (Table 1). Overall, 55% of participants were age 18-21 years, 63% were female, 48% were non-white and 16% were obese. Nearly half of participants were publicly insured, 14% were exposed to household smoking and the majority of parents had less than college education. Differences in prior season influenza vaccination (52% vs 47%) were not statistically significant (p=0.48).

**Table 1.**
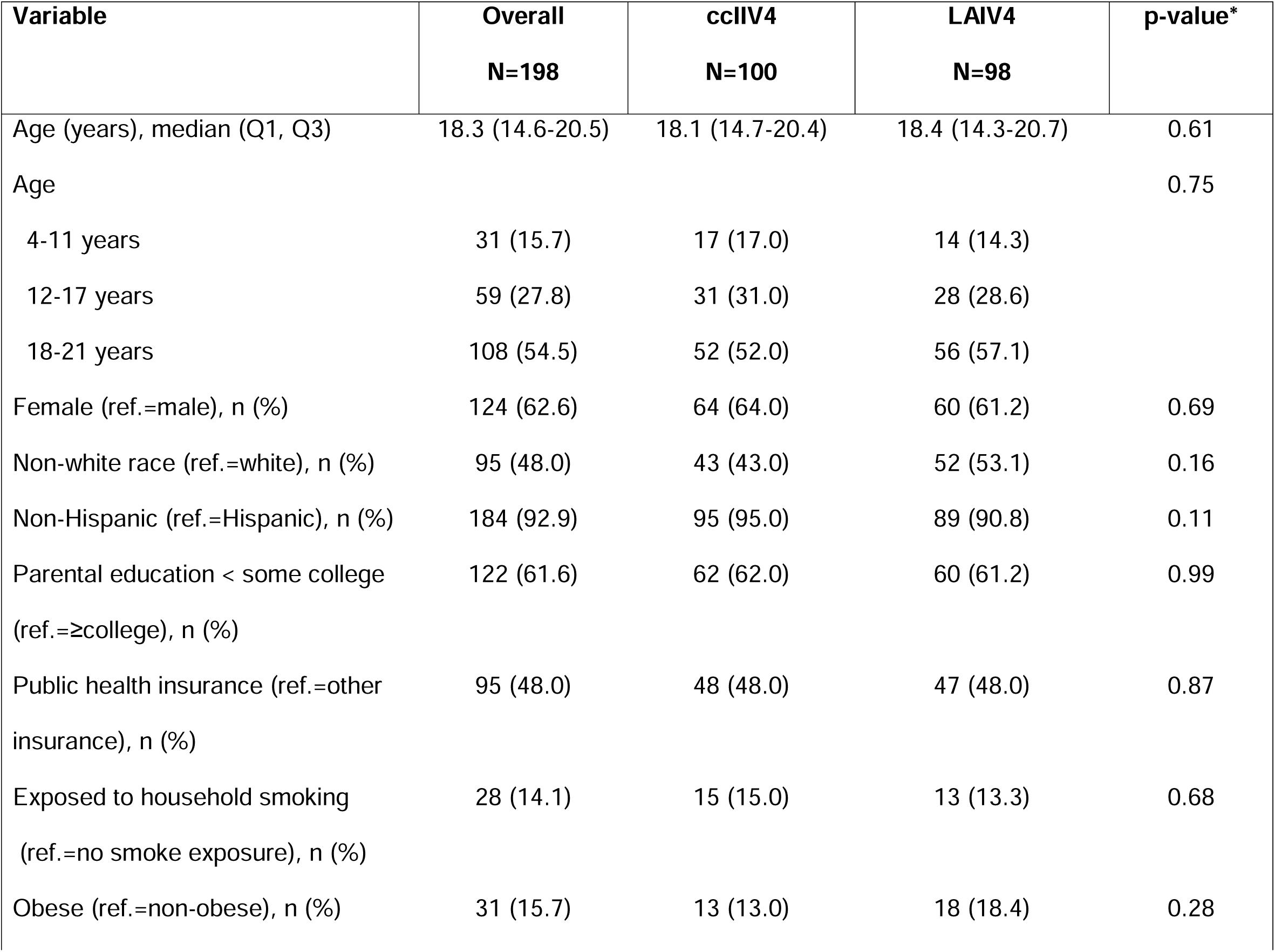
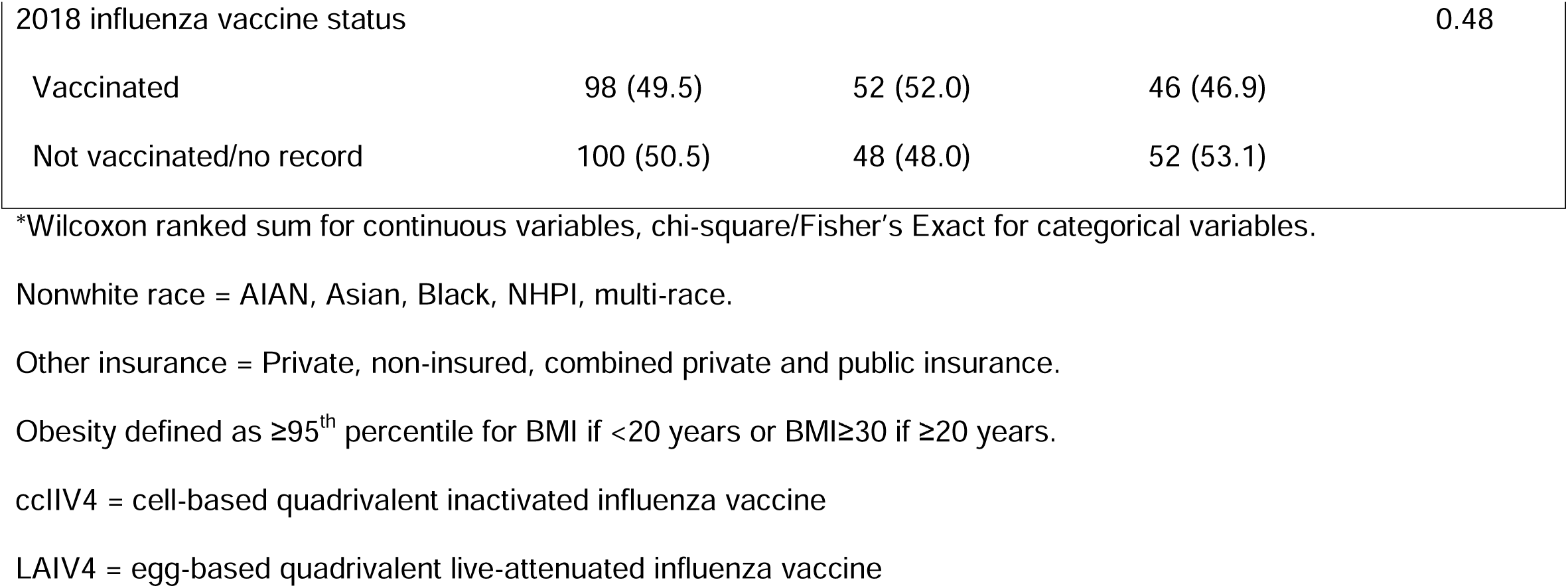
Demographics by 2019-20 influenza vaccine type received.

Pre- and post-vaccination GMTs and mean fold rise in HAI titers are shown by vaccine group in Table 2. Pre-vaccination titers were similar by vaccine group across all antigens; 72%-85% had titers ≥ 1:40 and 14%-61% had titers ≥ 1:110. After vaccination, GMTs increased significantly against all antigens except among LAIV4 recipients against B/Phuket (Yamagata lineage). All measures of post-vaccination titers, i.e., GMTs, GMFR and proportions with HAI titers ≥ 1:40 and ≥ 1:110, were significantly higher among ccIIV4 recipients than LAIV4 recipients across all antigens (all p≤0.02; Table 2). There was no significant difference between GMFR in antibody titer for A/H3N2 tested against cell-grown antigen and A/H3N2 tested against egg-grown antigen. Responses to the A/H3N2 antigens in ccIIV4 and LAIV4 recipients are shown in Figure 2.

**Table 2.** Pre- and post vaccination hemagglutination inhibition (HAI) assay titer responses by vaccine type.

|  | **cclIV4**<br>**N=100** | **LAIV4**<br>**N=98** | **p-value\*** |
| --- | --- | --- | --- |
| *A/H1N1-A/Brisbane Egg Grown Virus* |  |  |  |
| Day 0 HAI GMT (95% CI) | 90 (73-111) | 94 (77-116) | 0.79 |
| Day 28 HAI GMT (95% CI) | 269 (228-317)<sup>†</sup> | 108 (89-132)<sup>†</sup> | **<0.001** |
| Mean fold-rise in HAI titer (95% CI) | 3.0 (2.5-3.6) | 1.1 (1.0-1.3) | **<0.001** |
| Seroconversion ≥1:40, n (%) | 33 (33.0) | 5 (5.1) | **<0.001** |
| Day 0 HAI titer ≥1:40, n (%) | 83 (83.0) | 79 (80.6) | 0.66 |
| Day 28 HAI titer ≥1:40, n (%) | 96 (96.0) | 85 (86.7) | **0.02** |
| Day 0 HAI titer ≥1:110, n (%) | 61 (61.0) | 54 (55.1) | 0.40 |
| Day 28 HAI titer ≥1:110, n (%) | 93 (93.0) | 58 (59.2) | **<0.001** |
| *A/H3N2-A/Kansas Egg Grown Virus* |  |  |  |
| Day 0 HAI GMT (95% CI) | 78 (66-95) | 75 (64-89) | 0.79 |
| Day 28 HAI GMT (95% CI) | 205 (178-237)<sup>†</sup> | 96 (83-112)<sup>†</sup> | **<0.001** |
| Mean fold-rise in HAI titer (95% CI) | 2.6 (2.2-3.1) | 1.3 (1.1-1.4) | **<0.001** |
| Seroconversion ≥1:40, n (%) | 44 (44.0) | 11 (11.2) | **<0.001** |
| Day 0 HAI titer ≥1:40, n (%) | 85 (85.0) | 83 (84.0) | 0.95 |
| Day 28 HAI titer ≥1:40, n (%) | 99 (99.0) | 90 (91.8) | **0.02** |
| Day 0 HAI titer ≥1:110, n (%) | 41 (41.0) | 34 (34.7) | 0.36 |
| Day 28 HAI titer ≥1:110, n (%) | 77 (77.0) | 41 (41.8) | **<0.001** |
| *A/H3N2-A/Kansas Cell Grown Virus* |  |  |  |
| Day 0 HAI GMT (95% CI) | 46 (38-55) | 50 (43-58) | 0.56 |
| Day 28 HAI GMT (95% CI) | 136 (113-165)<sup>†</sup> | 63 (58-74)<sup>†</sup> | **<0.001** |
| Mean fold-rise in HAI titer (95% CI) | 3.0 (2.5-3.6) | 1.3 (1.1-1.4) | **<0.001** |
| Seroconversion ≥1:40, n (%) | 47 (47.0) | 8 (8.2) | **<0.001** |
| Day 0 HAI titer ≥1:40, n (%) | 72 (72.0) | 81 (82.7) | 0.07 |
| Day 28 HAI titer ≥1:40, n (%) | 93 (93.0) | 80 (81.6) | **0.02** |
| Day 0 HAI titer ≥1:110, n (%) | 24 (24.0) | 14 (14.3) | 0.08 |
| Day 28 HAI titer ≥1:110, n (%) | 65 (65.0) | 26 (26.5) | **<0.001** |
| *B/Victoria-B/Colorado Egg Grown Virus* |  |  |  |
| Day 0 HAI GMT (95% CI) | 65 (53-79) | 65 (53-81) | 0.95 |
| Day 28 HAI GMT (95% CI) | 158 (134-186)<sup>†</sup> | 78 (64-94)<sup>†</sup> | **<0.001** |
| Mean fold-rise in HAI titer (95% CI) | 2.4 (2.0-2.9) | 1.2 (1.0-1.4) | **<0.001** |
| Seroconversion ≥1:40, n (%) | 28 (28.0) | 9 (9.2) | **<0.001** |
| Day 0 HAI titer ≥1:40, n (%) | 75 (75.0) | 72 (73.5) | 0.81 |
| Day 28 HAI titer ≥1:40, n (%) | 96 (96.0) | 79 (80.6) | **<0.001** |
| Day 0 HAI titer ≥1:110, n (%) | 34 (34.0) | 34 (34.7) | 0.91 |
| Day 28 HAI titer ≥1:110, n (%) | 71 (71.0) | 36 (36.7) | **<0.001** |
| *B/Yamagata-B/Phuket Egg Grown Virus* |  |  |  |
| Day 0 HAI GMT (95% CI) | 89 (74-107) | 101 (83-122) | 0.44 |
| Day 28 HAI GMT (95% CI) | 215 (191-241)<sup>†</sup> | 107 (90-128) | **<0.001** |
| Mean fold-rise in HAI titer (95% CI) | 2.4 (2.0-2.9) | 1.1 (1.0-1.2) | **<0.001** |
| Seroconversion ≥1:40, n (%) | 29 (29.0) | 4 (4.1) | **<0.001** |
| Day 0 HAI titer ≥1:40, n (%) | 86 (86.0) | 84 (85.7) | 0.95 |
| Day 28 HAI titer $\geq 1:40$, n (%) | 100 (100) | 89 (90.8) | **0.002** |
| Day 0 HAI titer $\geq 1:110$, n (%) | 51 (51.0) | 57 (58.2) | 0.31 |
| Day 28 HAI titer $\geq 1:110$, n (%) | 84 (84.0) | 57 (58.2) | **<0.001** |

cclIV4 = cell-based quadrivalent inactivated influenza vaccine; LAIV4 = egg-based quadrivalent live-attenuated influenza vaccine; GMT = geometric mean titer; CI = confidence interval; Seroconversion = HAI titer ratio of day 28/day 0 $\geq 4$ and HAI titer at day 28 $\geq 40$ . Primary outcome was mean fold-rise in HAI titer.
\*t-test was used to compare log titers by vaccine type; Chi-square test was used to compare rates.
<sup>†</sup>p<0.05 for paired t-test comparison to Day 0 GMT.

**Figure 2.**
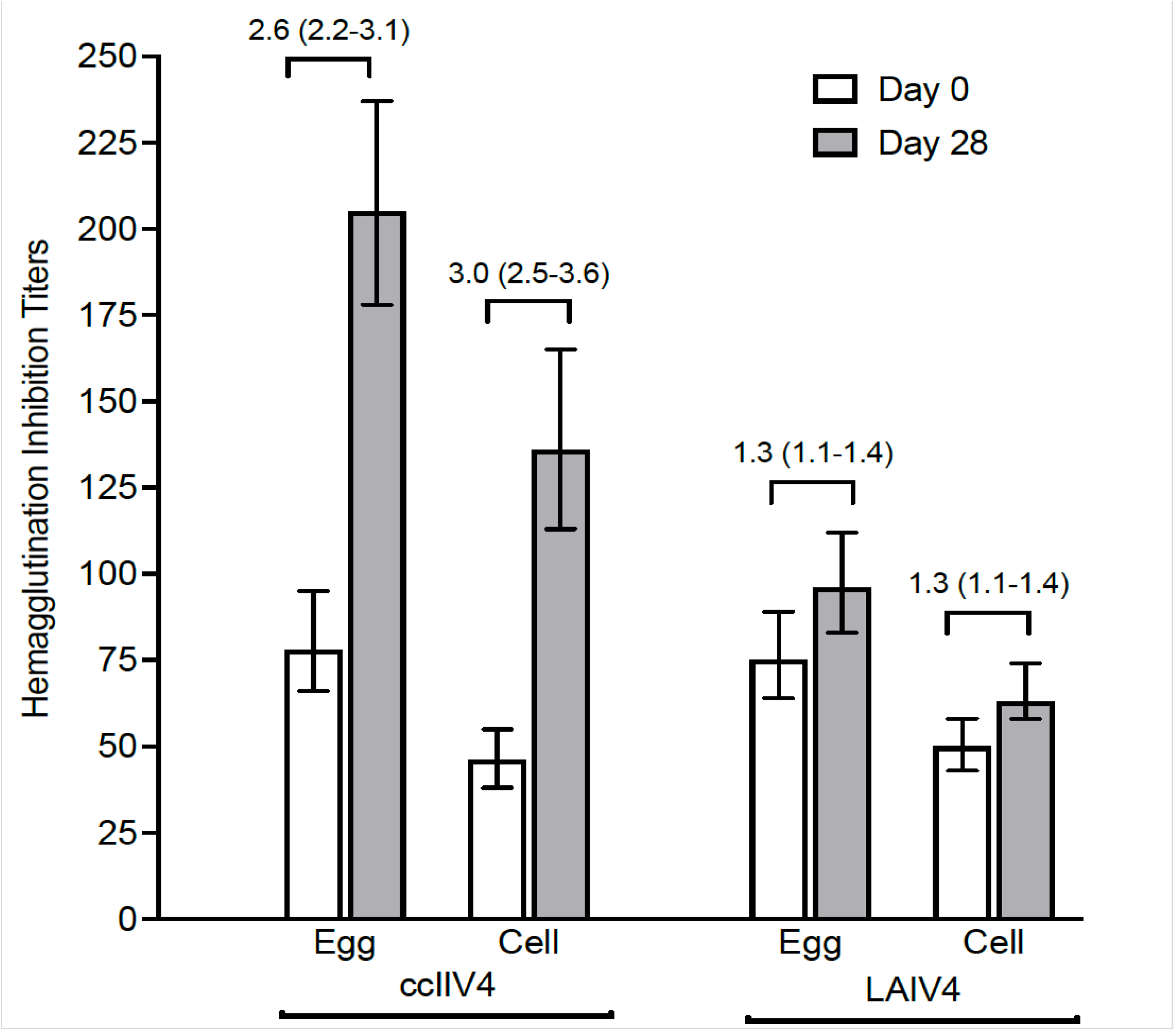
Pre- and post-vaccination GMT hemagglutination inhibition (HAI) assay titers by vaccine type for A/H3N2-A/Kansas egg and cell grown viruses. Data are day 0 (white) and day 28 (gray) with 95% confidence Intervals (CI), GMFR with 95% CI is shown horizontally above bars. Within each vaccine type, egg grown antigens were higher than cell grown at days 0 and 28 (p<0.001), but the GMFR did not differ between egg and cell. Between vaccine types, HAI titers at day 28 and GMFR were lower for LAIV4 for each respective antigen (p<0.0001).

Figures 3a and 3b show patterns of antibody response to vaccination for all antigens. For both ccIIV4 and LAIV4, antibody titers for most participants increased or remained similar to pre-vaccination; greater than two-fold decreased titers were observed in few participants in both vaccine groups. However, patterns of antibody response to vaccination differed significantly by vaccine type across all antigens, including against cell-grown and egg-grown A/H3N2 antigens. For the ccIIV4 recipients, a strong “ceiling” effect was observed, with post vaccination titers reaching similar levels across a range of prevaccination titers; whereas antibody titers among LAIV recipients increased more variably from pre- to post vaccination. In regression analyses, lower pre-vaccination titers were associated with greater GMFR in titers in both vaccine groups, although the association was much stronger in the ccIIV4 recipients. Controlling for pre-vaccination HAI titer, prior season (2018-19) vaccination was associated with decreased GMFR against B/Colorado-B/Victoria in ccIIV4 recipients (ß =-0.88, p<0.001). With similar adjustments, prior season vaccination was associated with increased GMFR against A/H3N2-A/Kansas egg-grown antigens (ß =0.17, p<0.01) in LAIV4 recipients, but was not independently associated with GMFR against other antigens.

**Figure 3a.**
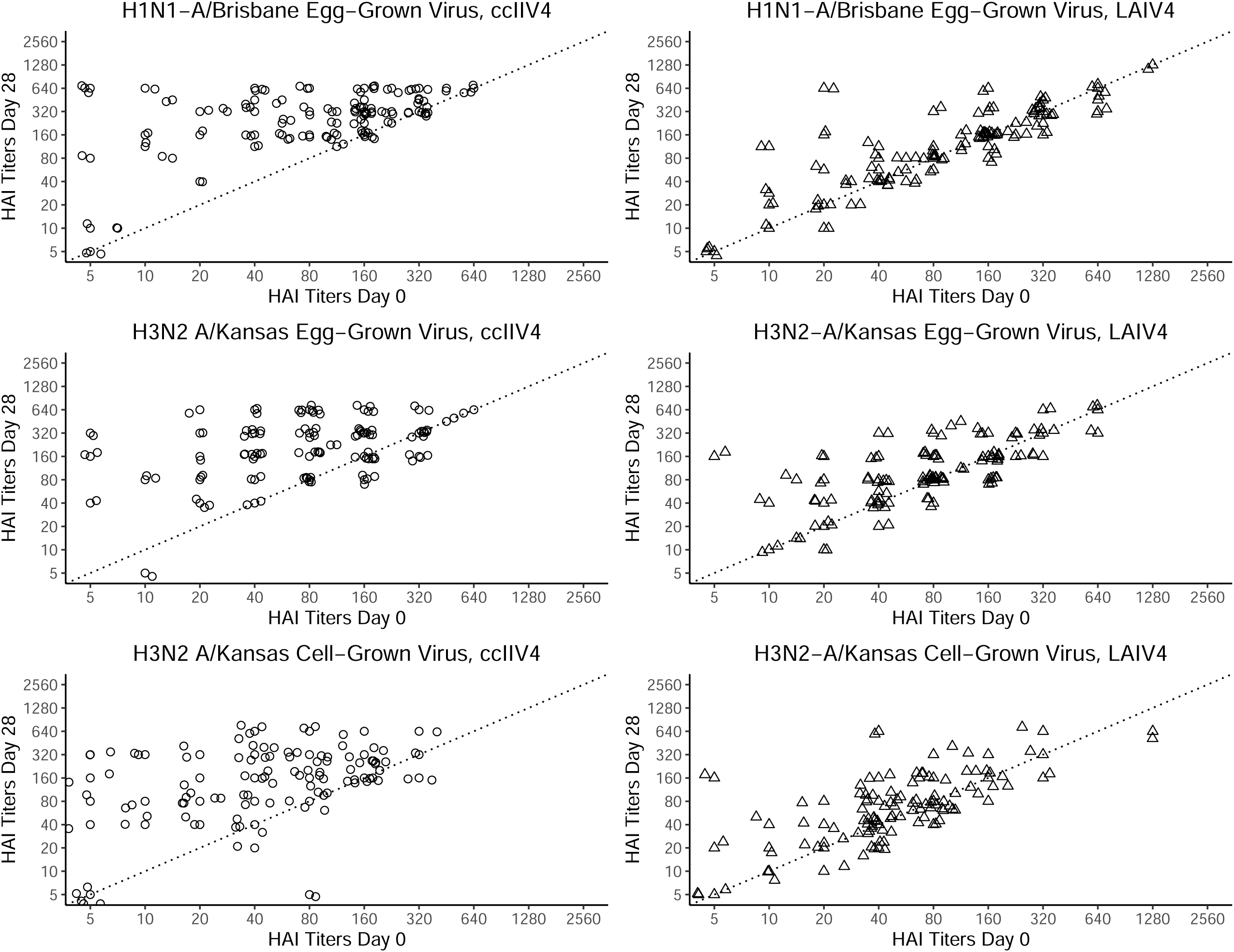
Hemagglutination inhibition (HAI) titers for influenza A antigens on day 0 and day 28 for ccIIV4 (left) and LAIV4 (right). Identical day 0 and day 28 titers fall on the dotted line. Data points above the line represent an increase in HAI at day 28 and data points below represent a decrease.

**Figure 3b.**
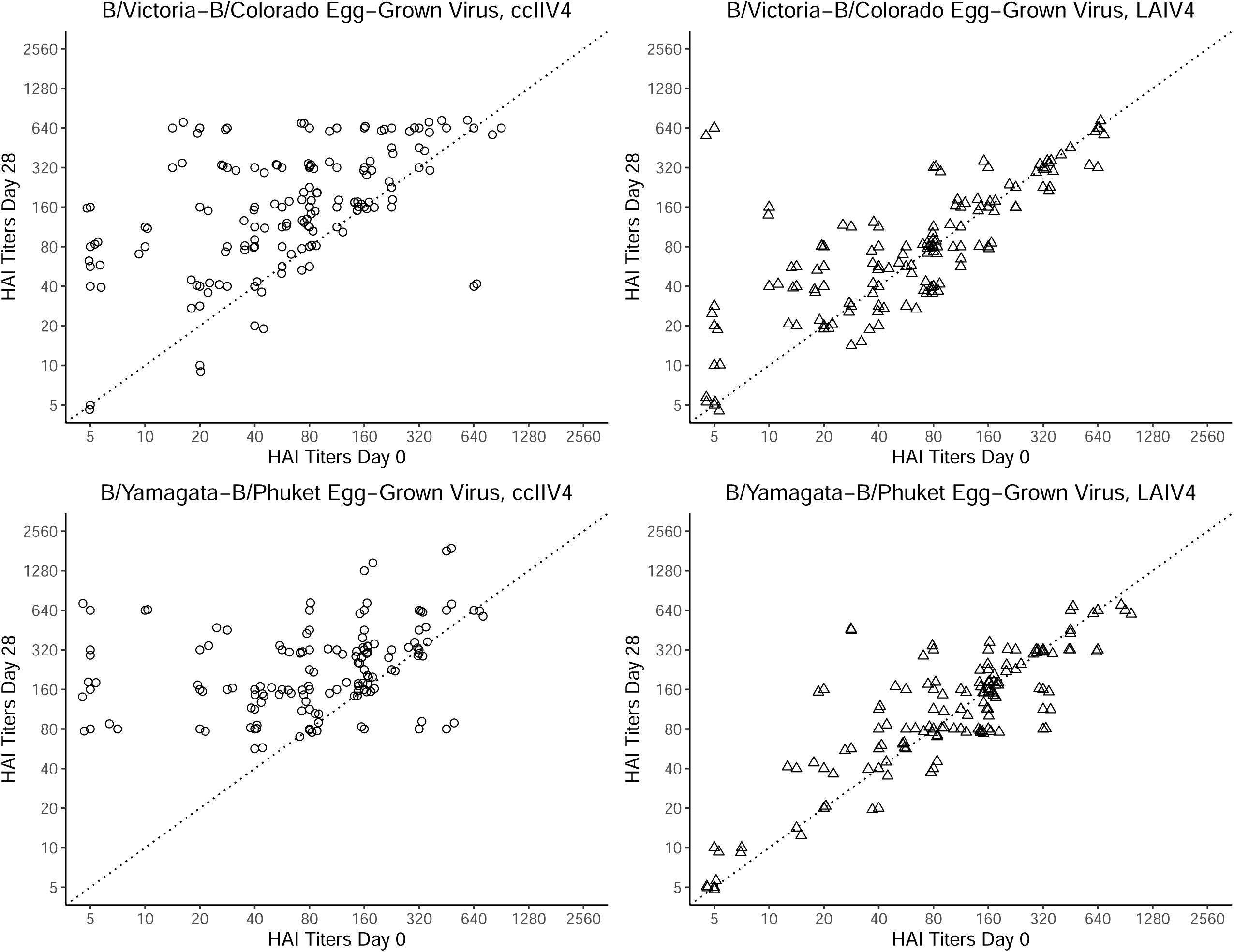
Hemagglutination inhibition (HAI) titers for influenza B antigens on day 0 and day 28 for ccIIV4 (left) and LAIV4 (right). Identical day 0 and day 28 titers fall on the dotted line. Data points above the line represent an increase in HAI at day 28 and data points below represent a decrease.

## 4. Discussion

This randomized controlled trial compared antibody responses to cell-grown (ccIIV4) and egg-grown (LAIV4) influenza vaccine in a racially diverse group of children and young adults. Compared to previous work, which used a ccIIV4 formulation with three cell-grown antigens and one egg-grown antigen (A/H1N1), this study used a ccIIV4 composed entirely of cell-based antigens. LAIV4 was composed entirely of egg-grown antigens. A/H3N2 strains have become increasingly glycosylated as a result of adaptations required for growth in eggs. These adaptations have led to antigenic differences between vaccine strains and the wild virus [15] and reduced vaccine effectiveness. Thus, cell-grown vaccine strains offer the potential to avoid HA protein mutations that occur in egg-based systems [16].

For all measures of HAI response, ccIIV4 elicited significantly larger increases than LAIV4 at 28 days post vaccination. For LAIV4, HAI response was attenuated and seroconversion was infrequent. Our findings are consistent with previous work showing .no HAI marker as a correlate of protection for LAIV4 [17].

Lower HAI responses to LAIV compared to IIV have been reported in prior seasons [3, 6, 9, 10]. Our findings with the 2019-20 LAIV4 formulation and fully cell-based ccIIV4 follow a similar pattern. Prior studies have similarly shown a lower HAI response to LAIV in countries where influenza vaccines are routinely administered, although a few earlier studies showed a significant HAI response to LAIV prior to the initiation of routine seasonal vaccination in children [17]. Because of different routes of administration for the two vaccine types, secretory IgA and cellular immune processes stimulated by administration via the nasal route may be a better measure of immune response than HAI for LAIV4 influenza vaccine [6, 10, 18]. Measures of immunoglobulins (IgG and IgA) and cellular immune responses would help delineate the potential for differences between the vaccine groups not observed in HAI titers alone.

When comparing the relationship of pre- to post vaccination titers, with ccIIV4 in particular, those with lower pre-vaccination titers had the greatest increase. For both ccIIV4 and LAIV4, participants with high baseline titers remained high. Pre-vaccination titer was predictor of post-vaccination titer (positively correlated) and GMFR (negatively correlated). We found similar relative GMFR increases in A/H3N2 cell- and egg-grown antigens within vaccine type in the 2019-20 season.

A prior study in 2018-19, comparing ccIIV4 with 3 cell-grown and 1 egg-grown strains and LAIV4, which incorporated microneutralization measures, found that the GMFR measured using egg-grown A/H3N2 antigen was higher for egg-based vaccine than cell-based vaccine recipients [12]. In the current study, GMFR responses to egg- and cell-grown A/H3N2 antigens were similar within vaccine type. Thus, the question of response based on the antigen growth medium is still unresolved.

Due to the known effect of baseline titers and prior vaccination history on vaccine response [18], we specifically recruited participants who were not vaccinated in the prior season anticipating that they may have lower titers pre-vaccination and be more likely to mount an HAI response. In post hoc analyses, pre-vaccination titer was a negative predictor of vaccine response for both vaccines, with a greater rise in titer among those with lower pre-vaccination titers. Despite the large proportion of participants unvaccinated in the prior year, only the HAI response to B/Colorado-B/Victoria components of both the 2018-19 and 2019-20 vaccines, had an independent association with a smaller GMFR. Prior year vaccination status predicted pre-vaccination titers and fold-rise in titers, vaccination with ccIIV4 remaining a stronger predictor of fold-rise after adjusting for prior year vaccination status. Pre-vaccination titers as well as prior year vaccination status should be considered in determination of degree of vaccine response.

### 4.1 Strengths and Limitations

A strength of this study was the relatively large randomized controlled sample. Small sample size within the youngest age group limited power to detect differences by age. Participants were historically highly vaccinated with high pre-vaccination titers, and repeated vaccination with egg-based vaccines may affect antibody responses to cell-based antigens. Overall, immune responses may not predict vaccine effectiveness, especially for LAIV for which better correlates of protection are needed.

### 4.2 Conclusions

In this racially diverse group of children and young adults, half of whom were unvaccinated in the prior season, all measures of HAI titers were higher among ccIIV4 recipients than LAIV recipients. Even among vaccine recipients with high pre-vaccination titers, HAI titers remained high post vaccination. Responses to ccIIV4 and LAIV4 as measured by GMFR were consistent against egg- and cell-grown antigens. Future studies are needed to examine the immunoglobulin and cellular immune responses in the 2019-20 season to ccIIV4 and LAIV4.

## Data Availability

Data can be made available upon request.

## Abbreviations

HAI: hemagglutination inhibition assay
IIV: inactivated influenza vaccine
ccIIV4: cell-culture-based inactivated influenza vaccine quadrivalent
LAIV4: Egg-based live attenuated influenza vaccine quadrivalent
EMR: Electronic medical record
RDE: Receptor-destroying enzyme
PBS: Phosphate-buffered saline
CDC: Centers for Disease Control and Prevention
FDA: Food and Drug Administration
GMT: Geometric mean titers
GMFR: Geometric mean fold rise
ACIP: Advisory Committee on Immunization Practice
PA-SIIS: Pennsylvania Statewide Immunization Information System

## Funding and support

This work was supported by the Centers for Disease Control and Prevention (CDC) [5U01IP001035] and by National Institutes of Health (NIH) [UL1TR001857], [KL2 TR001856], and/or [TL1 TR001858]. This work represents the views of the authors and not the CDC or NIH.

Pennsylvania Statewide Immunization Information System (PA-SIIS) vaccine registry was used to verify vaccination status. These data were supplied in part by the Bureau of Health Statistics & Registries, Pennsylvania Department of Health, Harrisburg, Pennsylvania. The Pennsylvania Department of Health specifically disclaims responsibility for any analyses, interpretations, or conclusions.

REDCap and the Department of Biomedical Informatics grant support (Clinical and Translational Sciences Institute at the University of Pittsburgh Grant Number UL1-TR-001857). Study data were collected and managed using REDCap electronic data capture tools hosted at the University of Pittsburgh.

## Conflict of Interest

RKZ has received funding by Sanofi for an unrelated study. MPN has research funding from Merck & Co., Inc. for an unrelated study. JMM has received funding from Merck, Sharp and Dohme for an unrelated study.

## Notes

### Clinical Trial

Clinical Trials No.: NCT03982069

